# Bayesian Multistate Modelling of Prefrailty and Frailty: A Microsimulation Study with Weight Management Interventions for Prevention in Asian Settings

**DOI:** 10.1101/2025.02.20.25322601

**Authors:** Xinyu Zhang, Shihui Jin, Haolong Song, Johnathan Yeo Hong-Hui, Borame Lee Dickens

## Abstract

**Background:** Pre-frailty and frailty prevalence is increasing in the Western Pacific, resulting in substantial morbidity and mortality in older populations. Modelling frameworks are required to estimate the prevalence of frailty and potential impacts of ongoing population-level nutritional interventions.

**Methods:** Using a microsimulation sociodemographic model of 3,353,032 individuals from 1990 to 2050, and data from the Singapore Longitudinal Ageing Study 2 of 3,270 participants, we developed a Bayesian multistate model of robust, pre-frailty and frailty stages with estimated transition probabilities by age, ethnicity, and gender for each body-mass index (BMI) category. We then explored four scenarios where weight management interventions are applied that modify the annual distribution of underweight, normal weight, overweight, obese I and obese II individuals.

**Findings:** Between 2011 and 2050, the overall prevalence of pre-frailty and frailty increased from 44·2% to 46.9%, and from 3·2% to 11·3% respectively. Reductions of 811 pre-frail individuals (95% CrI: 624-1,127) and 36,202 frail individuals (22,109- 41,124) were estimated when underweights shifted to normal weights, 5,787 (3,670- 7,707) and 55,777 (32,683-80,941) when obese II moved to obese I, and 22,045 (18,430-23,487) and 62,847 (40,165-91,517) when both groups shifted respectively. Total healthcare utilization decreases by 6·9% (4.3%-8·1%) with the latter intervention.

**Interpretation:** Frailty prevalence is projected to substantially increase by 2050 where large-scale weight management interventions could be utilised to avert cases of both pre-frailty and frailty in older individuals.

**Funding:** This research was supported by the Population Health Metrics and Analytics project, the Ministry of Health and National Innovation Challenge (NIC Ageing), Healthy Longevity Catalyst Awards (HLCA) MOH-HLCA22Feb-0007.

## Introduction

Frailty is defined as a clinically recognisable state, resulting from accumulative aging- associated decline across multiple physiologic systems, which compromises an individual’s ability to perform everyday tasks or cope with acute stressors. As a global public health concern among many countries’ ageing populations, frailty has been associated with increased hospitalization rates, functional decline, loss of independence, lower quality of life, and mortality^1,2^. A global systematic review of 21 studies reported that the prevalence of physical pre-frailty, a risk state which predisposes the development of frailty, ranged from 18·7%-53·1%, and frailty ranged from 4·0%-59·1% among community-dwelling older adults^3^. With populations continuing to age^4^, projections of frailty prevalence include 14·3% of women and 8·7% of men aged 60 or above by 2043 in Japan^5^, and an estimated doubling of prevalence for Norway^6^.

Within the Western Pacific, Singapore’s resident population is rapidly aging^7^. In 2024, the percentage of older persons aged 65 and over accounted for 13·7% of the population with the number exceeding 0·8 million individuals^8^. By 2050, the proportion is forecasted to reach 23·7%-30·4%, with the number exceeding 1·4 million^9^. The crude prevalence of frailty is therefore expected to rise although future estimates are scarce. Studies of community-living older adults in Singapore have estimated however that the prevalence of pre-frailty and frailty ranges from 40·1% to 44·2% and from 4·6% to 7·5%, respectively, up until 2013^10,11,12^. The increasing prevalence of frailty poses significant challenges on healthcare systems^4,13,14^ where those who are underweight or obese, already facing increased likelihoods of complications from injury^15,16^ or chronic diseases^17,18^, are at greater risk^19^.

A meta-analysis of 17 studies including 78,642 community-dwelling older adults from 10 countries in America, Europe, and Asia found that obesity (body mass index (BMI) ≥ 30 *kg*/*m*^2^) and underweight (BMI < 18·5 *kg*/*m*^2^) were associated with significant higher risks of frailty (relative risk (RR) = 1·40, 95% CI: 1·17–1·67 for obesity and RR = 1·45, 95% CI: 1·10–1·90 for underweight, respectively), and a U-shaped relationship existed between BMI and the risk for frailty with a nadir of risk at a BMI of 18·5–29·9 *kg*/*m*^2^ ^19^. Obesity in particular^19^, which is becoming increasingly prevalent among older adults as sarcopenic obesity^20^, has been linked with frailty in several cross-sectional and prospective population-based studies where a negative relationship has been described^21–25^. The direct mechanisms include increased joint stress and metabolic inflammation, which reduces mobility and leads to the loss of muscle mass and strength. In terms of interventions, a literature review including 21 studies from the United States, Australia, Canada, and Italy identified strong clinical evidence that weight loss and exercise interventions were effective in improving physical function among obese frail older adults^26^. For underweight older adults, multiple strategies have been taken to prevent weight loss and frailty, and to increase body weight, including supplementing diets with protein, energy, and vitamins in the US^27^ and France^28^.

In Singapore, efforts are underway to decrease frailty rates through multiple weight management and physical health schemes, targeting older adults^29^. A key nutritional intervention is a set of community-based meal delivery schemes, Meals-On-Wheels, aiming to address malnutrition and undernutrition among older adults who may have difficulties in preparing meals at home or accessing local food establishments that may not serve their nutritional needs. The provisioning of two meals per day, which are nutritionally balanced, encouraging healthy eating habits, can support special dietary needs and allow for regular check-ins to monitor the nutritional^30^ status of individuals, and can be utilized for weight management. In such programmes, studies^31,32^ have shown that significant reductions can be observed in overall weight and BMI among those who are overweight, and increases in protein intake and weight gain occurred after meal supplementation among those who are malnourished^33,34,35^. Through these intervention groups, the transitioning of underweights to normal weights, and obese to less obese weight categories represents key goals for healthy ageing, but the impact of these on future pre-frailty and frailty prevalence is still unknown. Such interventions are only explored in very small-scale studies, which requires exploration in large-scale models in order to support decision-making for intervention deployment and potential scale-up.

Our study therefore aims to explore the impact of weight management interventions on projections of pre-frailty and frailty utilising Singapore as a case study. To estimate the prevalence of pre-frailty and frailty, and explore the impact of interventions on BMI, we built a microsimulation model for 3,353,032 adults aged 55 and above across three main ethnicities—Chinese, Malays and Indians, and for men and women. We calculated transition probabilities between different frailty states using a Bayesian multistate model that accounted for estimated BMI trajectories across each individual’s lifetime to where we modelled three main scenarios examining the change in pre-frailty and frailty prevalence from reducing the proportion of older adults that were underweight, very obese and both. In then comparing potential reductions of frailty, evidence-based targeting of interventions can be supported.

## Methods

We built a microsimulation population model to test the efficacy of interventions designed to reduce frailty among older individuals from 2011 to 2050. We assigned frailty states to individuals from 2011 to 2016 based on data from the Singapore Longitudinal Ageing Study 2 and literature^10,11^. We then built a Bayesian multistate model to calculate transition probabilities by age, ethnicity, and gender and applied the transition probabilities to individuals across time to obtain nationwide projections of prefrailty and frailty prevalence from 2011 to 2050. Additionally, we explored the effectiveness of three weight management interventions in modifying estimated prevalences.

### DEMOS Model

We used the Demographic Epidemiological Model of Singapore (DEMOS) to generate individual data, representing each resident in Singapore from 1990 to 2050^36^. The DEMOS leveraged fertility rates, mortality rates, migration, and BMI trajectories from sub-models to project the resident population of Singapore each year. We forecasted ethnicity-specific fertility rates using Bayesian Structural Time Series (BSTS) models by MCMC. We applied a logarithmic transformation to account for the decelerating decline in fertility rates and used annual fertility rates by ethnicity^37^ during periods when fertility rates fell below 2·1 for two consecutive years^38^. Annual Age-specific mortality rates from 1990–2023 were obtained from the Singapore Department of Statistics^39^, and mortality rates from 2023–2050 were projected using the Lee-Carter model with logarithmic transformation applied to age-specific mortality rates from 2010–2019. We found constant migration rates and age distribution of migrants by comparing simulated and actual populations, and assumed the stability in migration patterns for the projection period. The hierarchical model of BMI trajectories, stratified by gender and ethnicity, described each individual’s BMI over time as Gaussian fluctuations around a sequence of connected lines, with model parameters estimated using statistics from the Singapore Department of Statistics and national surveys^40^.

The population projections were validated by comparing with the population structure of each one-year age group, gender and ethnicity stratum released by the Singapore Department of Statistics from 1990 to 2023^41^. The projections of BMI were checked by comparing the projected proportions of four BMI categories (underweight, normal weight, overweight, and obese) with proportions observed in the 2020 and 2022 National Population Health Surveys^42^.

### Identification of Frailty Status

We adopted the Fried’s phenotype criteria which focus on physical frailty^43^ and assesses frailty using five parameters: weakness, slowness, shrinking (unintentional weight loss), exhaustion, and low physical activity. A test-taker is defined as robust if meeting none of the parameters, as pre-frail if meeting one or two parameters, and as frail if meeting three or more parameters^10^. With individual data generated by the DEMOS from 1990 to 2050, we calculated the total numbers of robust individuals, pre-frail individuals, and frail individuals in 2011 and 2016 based on the prevalence of frailty states among Singapore older residents aged 55 years and over from the Singapore Longitudinal Ageing Study 2 (SLAS-2) in 2011 and a 5-year follow-up in 2016^10^, which reported the prevalence of pre-frailty and frailty in 2011 as 44·2% and 3·4%, and as 48·9% and 7·5% in 2016.

Based on the proportions of the robust, pre-frail and frail individuals among older adults across age (55 to 100 years), gender (male, female) and ethnicity (three main ethnicities—Chinese, Malay and Indian), collected from a cross-sectional survey with a nationally representative sample of 2102 Singapore residents using Fried’s phenotype^11^, we estimated the number of individuals in each strata for the states of robust, prefrailty and frailty between 2011 and 2016. We used Bernoulli trials based on transition rates derived from prefrailty and frailty prevalences in 2011 and 2016 to simulate individual transitions between robust, pre-frail, and frail states from 2011 to 2016.

### Bayesian Multistate Model

We built a Bayesian multistate model to estimate annual transition probabilities differing for each age, ethnicity, and gender. Our model was built on the mechanism of a Bayesian three-state model which defined the binomial likelihoods and priors for the observed cross-sectional data and unknown parameters given by the disease incidence rate, the remission rate, and the case fatality rate^44^. Nonlinear spline functions of age were used with Hamiltonian MCMC to approximate the posterior distributions of the rates. The formulars and schematic presentation of the Bayesian multistate model are provided in Supplementary Material for clarity. We assumed that a robust individual may stay robust or transition to pre-frailty, a pre-frail individual may revert to robust, stay pre-frail or transition to frail, and a frail individual may revert to pre-frail or stay frail in one year. Annual incidence and remission probabilities between robust, pre-frailty, and frailty states were fitted. We did not however explicitly model frailty- induced mortality. We calculated five-year age-ethnicity-gender specific transition probabilities from robust to pre-frailty, from pre-frailty to frailty, and the remission probabilities from frailty to pre-frailty, and from pre-frailty to robust for older adults at each age from 55 to 85, and for age groups 86–87, 88–90, and >90, and then transformed them to annual transition probabilities by the formula *P*_1_ = 1 − (1 − *P*_5_)^1/5^ where *P*_*x*_ represents the transition probability for duration *x* years.

### Validation of Estimated Prevalence

We applied the annual transition probabilities to each individual aged 55 or over in DEMOS and generated their states in the next year using a Bernoulli distribution or categorical distribution based on their assigned states in 2011. We repeated such steps and projected the states for each individual in the following five years until 2016. Validation checks were performed by comparing the projected prevalence of pre-frailty and frailty in 2016 for older adults aged 60 and over in DEMOS with the observed prevalence for community-dwelling older Singaporeans aged 60 and over in a 5-year follow-up in the SLAS-2^10^.

### BMI Intervention Modelling

To explore the association of BMI on frailty states in Singapore, we simulated BMI and frailty states based on the characteristics of participants in national surveys in Singapore^10,11^. We classified BMI values into five categories in accordance with the Asia-Pacific classification: underweight (<18·5 *kg*/*m*^2^), normal weight (18·5–22·9 *kg*/*m*^2^), overweight (23·0–24·9 *kg*/*m*^2^), obesity I (25·0–29·9 *kg*/*m*^2^), and obesity II (≥30·0 *kg*/*m*^2^)^45^. We used two logistic regression models to fit the simulated data and calculated the odds ratios of pre-frailty and frailty, respectively, for each BMI category relative to the reference category (normal weight). We found that the majority of individuals had a normal weight in DEMOS (34% proportion of individuals), so the original transition probabilities can be treated as those for the normal weight, the reference BMI category. We derived the incidence rate for the normal weight BMI category *I*_(*inc,i*)_based on a continuous-time Markov model^46^

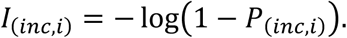

Then we applied the original incidence rate between the frailty states to the reference BMI category and estimated the incidence rates of other BMI categories using the formula *I*_(*inc,i*)_ = *OR*_*i*_ ⋅ *I*_(*i*𝑛𝑐,𝑟)_based on previously established methods^47–49^ which demonstrate that odds ratios can be used to estimate incidence ratios. *I*_(*inc,i*)_ denotes the incidence rate of pre-frailty or frailty for the BMI category *i*, *OR*_*i*_ denotes the odds ratio for the BMI category *i* relative to the reference category, and *I*_(*inc,r*)_denote the incidence rate of transitioning to pre-frailty or frailty states for the reference BMI category. The approximate remission probabilities were calculated by similar formulas, but the odds ratio *OR*_*i*_ was replaced by its inverse. Then we derive the annual incidence probability *P*_(*inc,i*)_using the previous continuous-time Markov model^46^

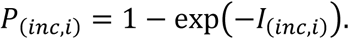

### Addition of Frailty-Related Mortality

We utilised a cohort study of 10,912 adults in the Kyoto-Kameoka Study in Japan^50^ to determine a hazard function for mortality based on frailty status and BMI. This study estimated a multivariable-adjusted Hazard Ratio (HR) between non-frail normal weight (BMI: 21.5 to 24.9) and other frail status BMI categories, adjusting for age and sex. Thus, we derived the frailty-related mortality rate for each BMI *i* category based on the original mortality rate *P*_(*m*𝑜𝑟𝑡𝑎𝑙*i*𝑡𝑦,𝑟)_for robust individuals in the reference group and corresponding hazard ratio 𝐻𝑅_*i*_ using the previous continuous-time Markov model^46^

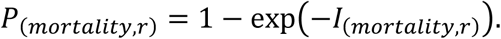

Then the mortality rates of pre-frail and frailty individuals in the BMI category *i* are estimated by

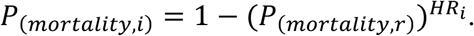

We assumed that the hazard ratios of pre-frailty were half of those observed of frailty based on a systematic review of 24 studies^51^.

### Scenarios

We considered four scenarios based on the established relationship between frailty and BMI: the Baseline Scenario without a weight management intervention where all individuals’ BMI categories remained unaltered in their estimated trajectories; the Underweight Scenario where a proportion (𝑝) of underweight individuals were shifted to normal weight; the Obese II Scenario (Obese II) where 𝑝 of all obese II individuals were shifted to obese I; the Both Scenario (Both) where 𝑝 of underweight individuals were shifted to normal weight and 𝑝 of obese II individuals were shifted to obese I. Here, we set the parameter 𝑝 to 5%, 10%, 25%, 50%, and 100%, representing low, moderate, and high effectiveness of weight-control interventions, respectively.

### Model Output

Based on the assigned frailty states in DEMOS in 2011, we projected pre-frailty and frailty prevalence across each stratum at the Baseline Scenario. From 2024 onwards, we adjusted BMI values according to the assumptions of the three scenarios with weight management interventions and projected pre-frailty and frailty numbers and prevalence for each scenario. We applied 12 groups of transition probabilities within their 95% credible intervals to the individuals in DEMOS by implementing 30 projections for each group to obtain uncertainty intervals for each scenario. All data analyses were performed by R software (version 4·4·1).

### Estimation of Healthcare Utilization

To further illustrate the effectiveness of weight management interventions on reducing frailty and associated healthcare costs, we estimated healthcare utilization in different care settings including specialist outpatient clinic visits (SOCV), emergency department visits (EDV), and hospitalizations. We referred to the healthcare utilization data of community-dwelling older adults aged 60 and over, collected from the Population Health Index study covering two periods: six months before and six months after the observation of frailty states^14^. We estimated healthcare utilization for each year by multiplying the number of robust, prefrail, and frail individuals by the mean number of SOCV, EDV, and hospitalizations expected for each state within a year, which was the sum of the means collected six months before and after the observation, respectively. The six-month means of healthcare utilization for each state are provided in Supplementary Material (Table S1).

## Results

### Transition Probabilities

Annual incidence and remission probabilities between robust, pre-frailty, and frailty states increase and decrease, respectively, with age across all ethnicities and genders (Figure 1). For Chinese men, the incidence probability of transitioning from robust to pre-frailty rises from 0·063 (95% CrI: 0·052–0·074) at age 55 to 0·242 (0·178–0·309) for those older than 90. Chinese women showed a similar trend, but with higher probabilities, reaching 0·329 (0·290–0·362) for individuals aged over 90. For Malays and Indians, these probabilities are 2·2%–4·8% and 1·9%–5·3% higher, respectively, than for Chinese. Incidence probabilities between pre-frailty and frailty followed similar age-related trends but are 4·2%–27·9% lower and increase more gradually.

**Figure 1:**
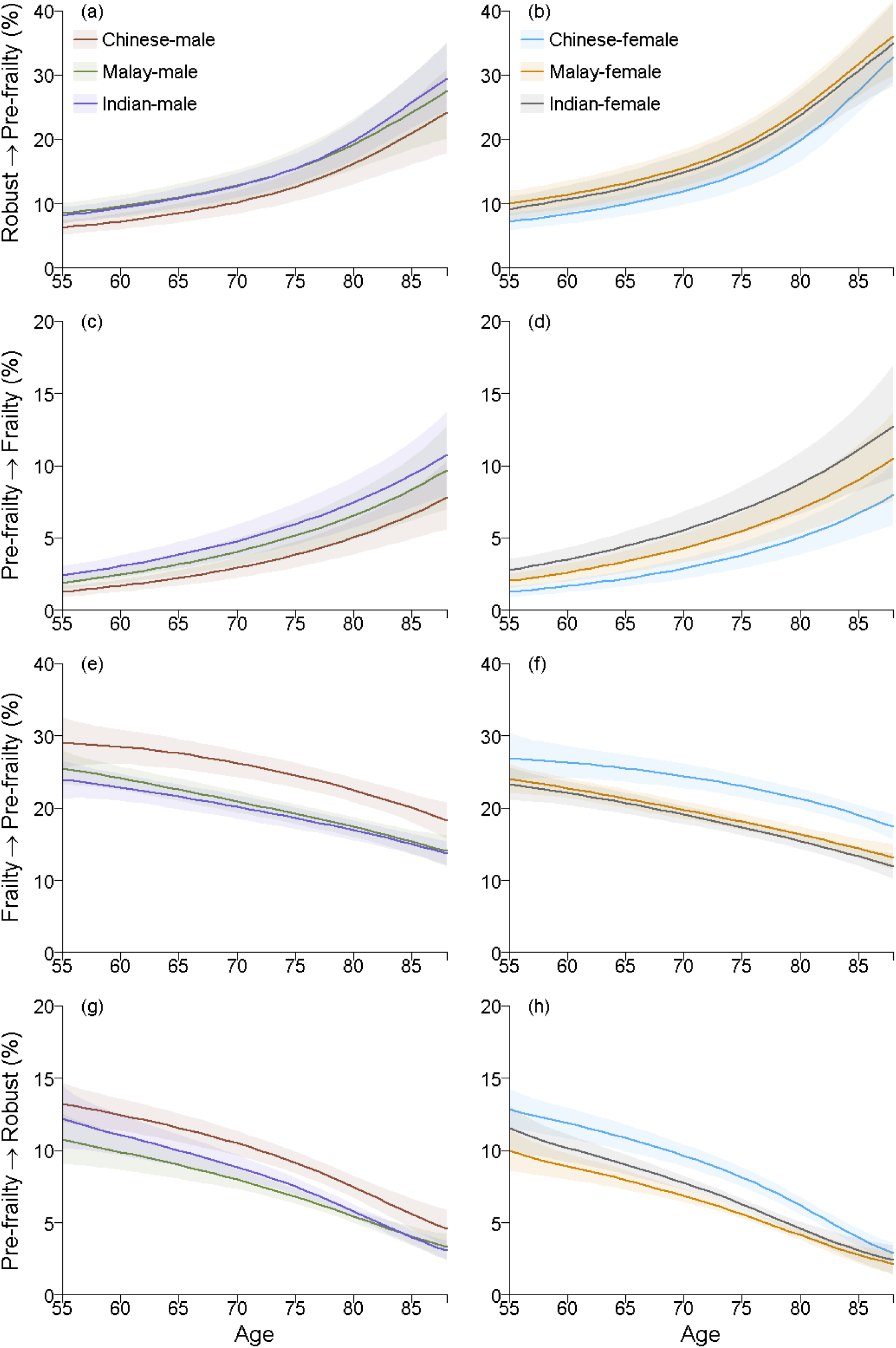
The annual transition probabilities (posterior means and 95% credible intervals as shaded areas) stratified by ethnicity (Chinese, Indian, and Malay). (a) robust to pre-frail men, (b) robust to pre-frail women, (c) pre-frail to frail men, (d) pre-frail to frail women, (e) frail to pre-frail men, (f) frail to pre-frail women, (g) pre-frail to robust men, (h) pre-frail to robust women.

Remission probabilities between frailty and pre-frailty decrease with age, from 0·290 (95% CrI: 0·255–0·325) to 0.183 (0·157–0·208) for Chinese men. For Chinese women, these probabilities are lower, declining from 0·269 (0·237–0·304) to 0·174 (0·157– 0·192). Malays and Indians have 2·9%-5·4% and 3·0-6·1% lower remission probabilities, respectively. Similarly, remission probabilities between pre-frailty and robust decline with age but are 8·8%–17·8% higher than those between frailty and pre-frailty.

### Projection Results across Weight Management Scenarios

In the Baseline Scenario without weight management interventions, the projected pre-frailty prevalence increased from 44·2% to 46.9% (95% CrI: 45·7%–47·5%), and frailty prevalence rose from 3·2% to 11·3% (7.1%–14.4%) by 2050 (Figure 2). The total numbers of pre-frail and frail individuals grew to 836,117 (792,202–860,643) and 201,859 (122,245–263,199), respectively (Figure 2), which are 2.3 and 7.8 times higher than those in 2011.

**Figure 2:**
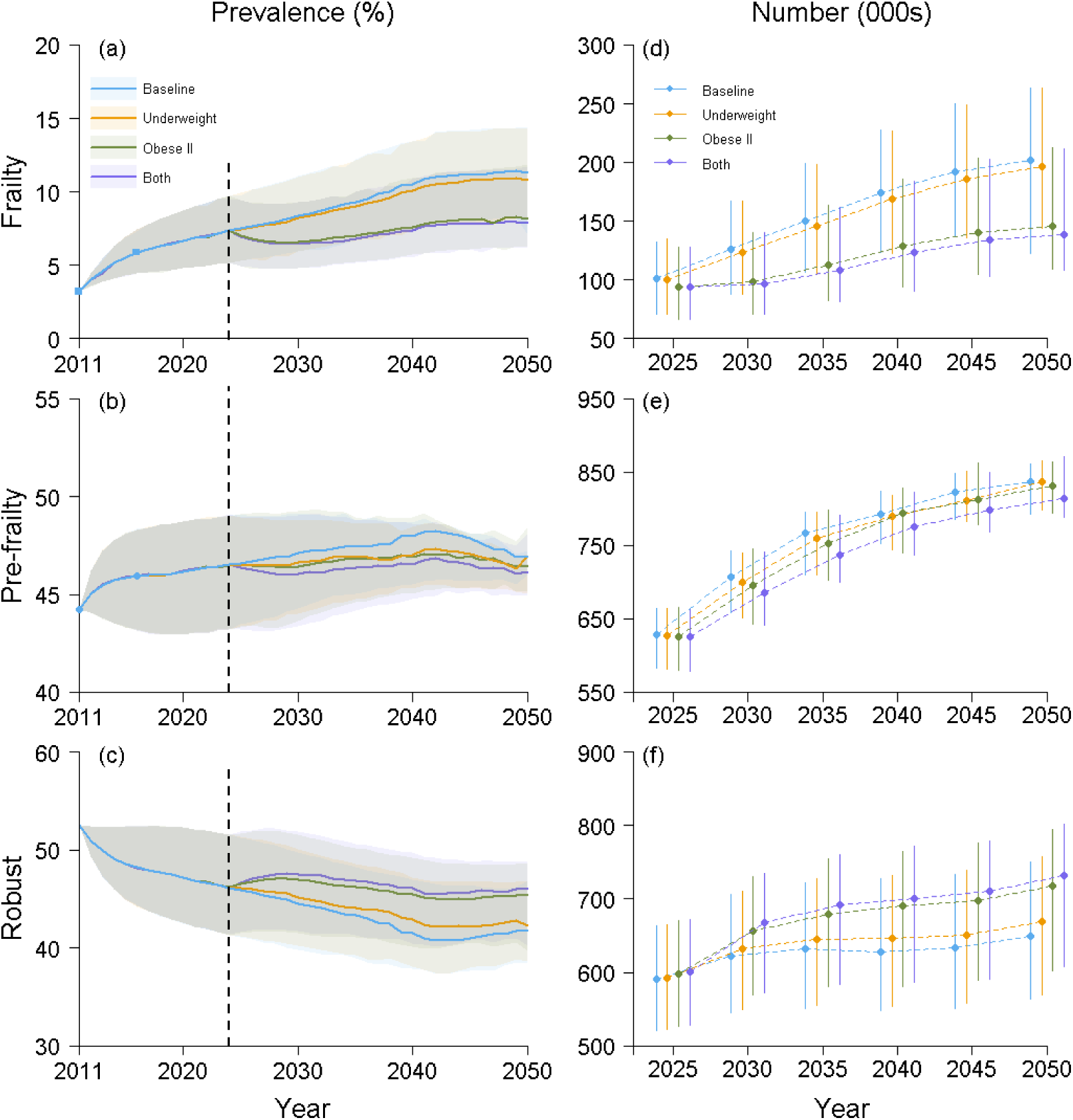
Projected prevalences and numbers of the frail, pre-frail, and robust individuals (posterior means and 95% credible intervals as shaded areas and error bars) for the whole population under four scenarios: baseline, underweight to normal weight intervention (“Underweight”), obese II to obese I intervention (“Obese II”), and the combination of both Underweight and Obese II strategies (“Both”). (a) frail prevalence, (b) pre-frail prevalence, (c) robust prevalence, (d) numbers of frail individuals, (e) numbers of pre-frail individuals, (f) numbers of robust individuals.

By ethnicity, in 2050, Chinese men had the lowest projected prevalence of pre-frailty (43·8%, 95% CrI: 41·5%–44.7%) and frailty (8·1%, 95% CrI: 5·0%–10·7%), while Chinese women had slightly higher prevalences (49·3%, 95% CrI: 48.3%–49.8%; and 9·5%, 95% CrI: 5.8%–12.4%) (Figure S2). For Malays, pre-frailty and frailty prevalences are 1·00%–9.1% and 91.5%-142.5% higher than those for Chinese. For Indians, pre-frailty prevalence is 5·2% higher for men (0.7%-15·4%) but 10.0% lower for women (1·0%–15·1%) than Chinese. Frailty prevalences among Indians exceed those of Chinese by 128.7%–227.3%.

With weight management interventions starting in 2024, the Both Scenario reduces the number of pre-frail and frail individuals by 22,045 (95% CrI: 18,430-23,487) and 62,847 (40,165-91,517), respectively, by 2050, corresponding to a decrease in pre-frailty and frailty prevalences by 2·6% (0·5%–3.2%) and 31.1% (19·6%–40.5%) compared to the Baseline Scenario. The Underweight and Obese II Scenarios individually also reduce pre-frail individuals by 811 (624-1,127) and 5,787 (3,670-7,707), and frail individuals by 36,202 (22,109-41,124) and 55,777 (32,683-80,941), respectively, compared to the Baseline Scenario, though these reductions are smaller than those in the Both Scenario.

### Delay in Transition of Pre-frailty and Frailty

We evaluated the effectiveness of BMI interventions by comparing differences in the numbers of new frail individuals, remaining robust individuals, and new robust individuals between scenarios with weight management interventions and the Baseline Scenario from 2025 to 2050 (Figure 3), and the changes in new and total frail, pre-frail, and robust individuals in age group composition in 2030 and 2050 (Figure 4). Five cases were considered in Figure 3, where interventions are effective for 100%, 50%, 25%, 10%, and 5% of target individuals.

**Figure 3:**
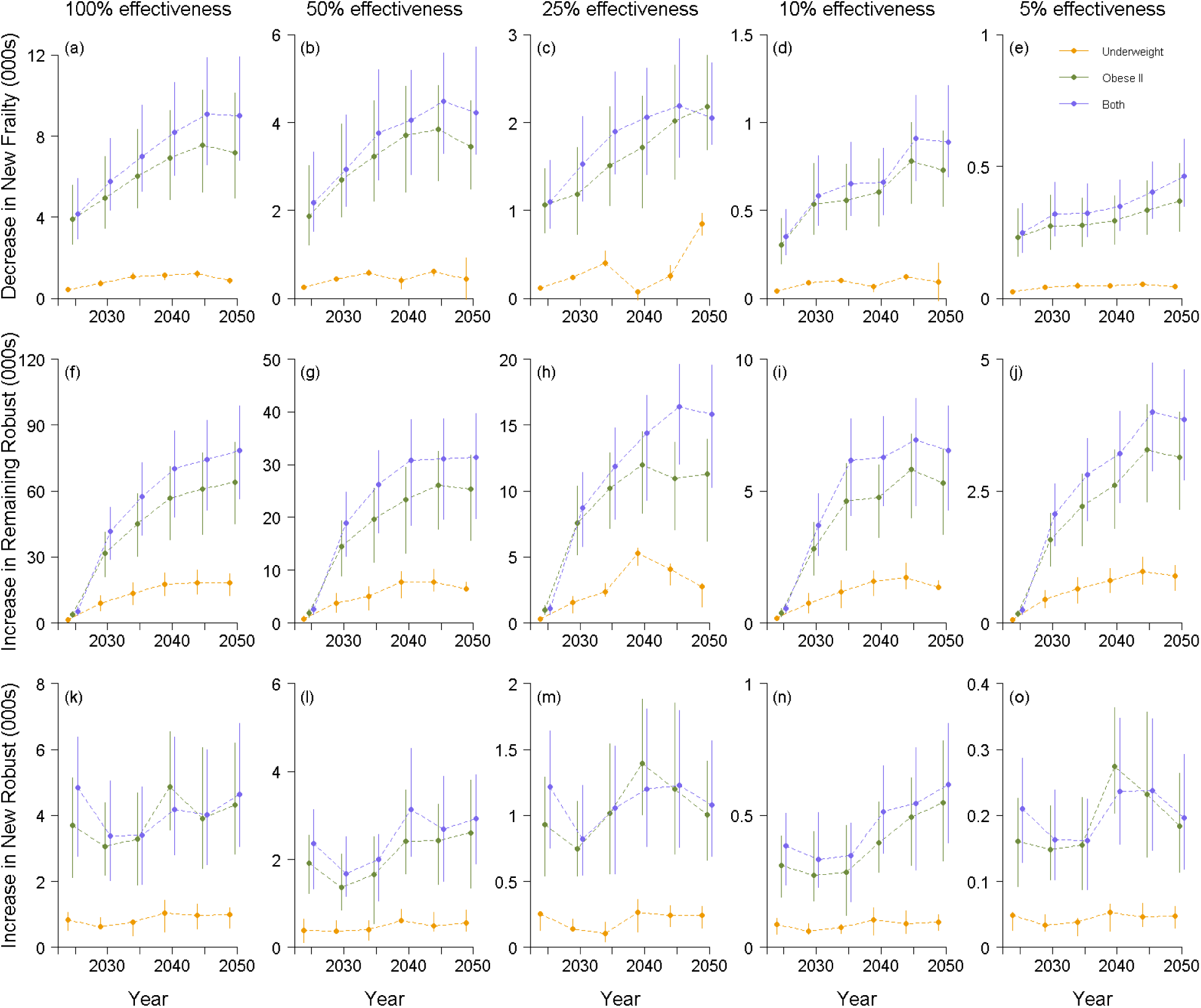
The difference between the numbers of new frail individuals (Row 1), remaining robust individuals (Row 2), and new robust individuals (Row 3) in each scenario with weight management interventions (Underweight, Obese II, Both) and the Baseline scenario from 2025 to 2050. The five columns represent five levels of weight-control intervention effectiveness, with 100%, 50%, 25%, 10%, and 5% of the obese II individuals shifting to obese I or underweight individuals shifting to the normal weight category. Estimates are presented as **posterior means and 95% credible intervals as error bars**.

**Figure 4:**
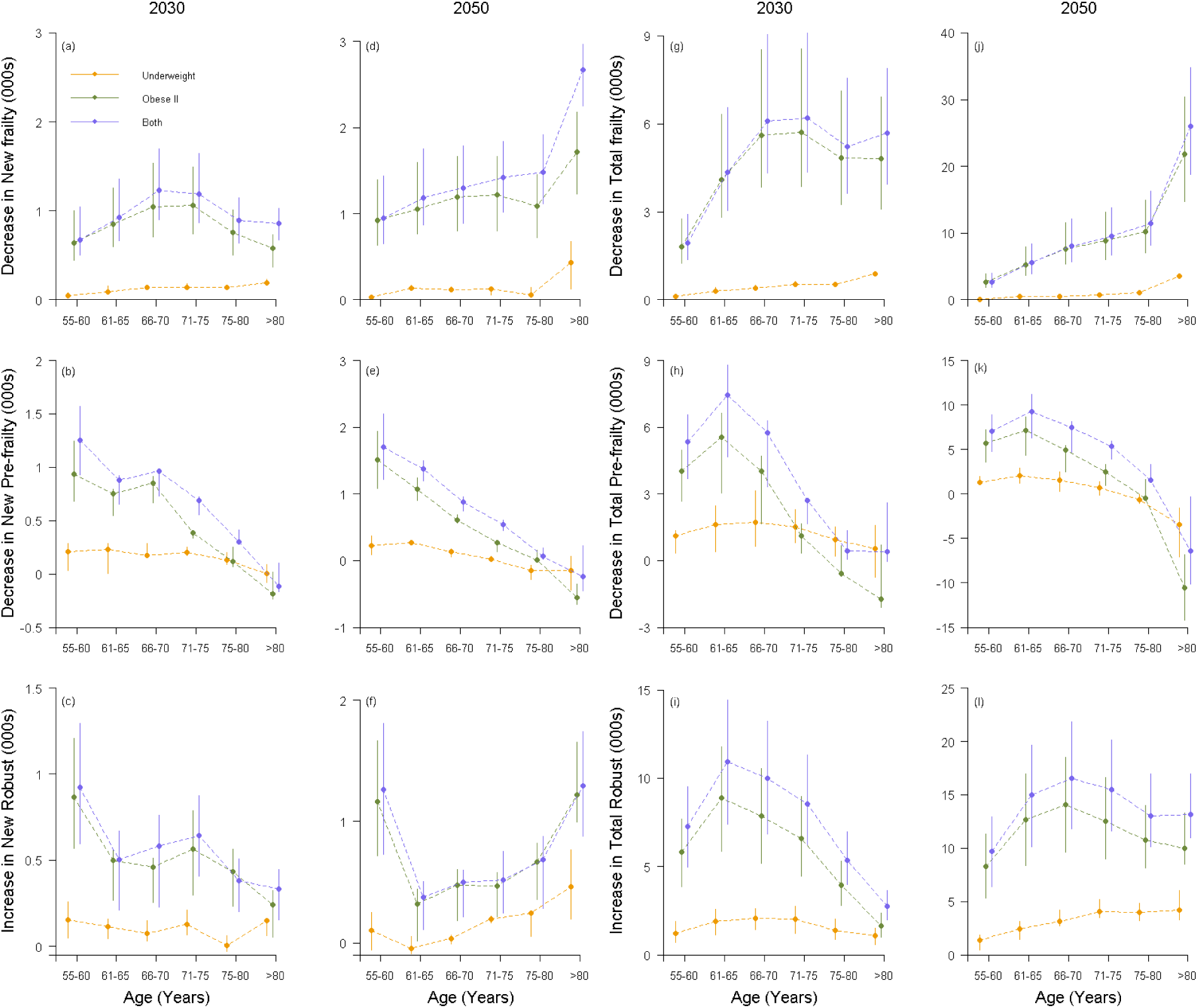
Changes in the numbers of new and total frail, pre-frail, and robust individuals (posterior means and 95% credible intervals as error bars) across age groups under each scenario with weight management interventions (Underweight, Obese II, Both) and the Baseline scenario in 2030 and 2050. (a) decrease in new frail individuals in 2030, (b) decrease in new pre-frail individuals in 2030, (c) increase in new robust individuals in 2030, (d) decrease in new frail individuals in 2050, (e) decrease in new pre-frail individuals in 2050, (f) increase in new robust individuals in 2050, (g) decrease in total frail individuals in 2030, (h) decrease in total pre-frail individuals in 2030, (i) increase in total robust individuals in 2030, (j) decrease in total frail individuals in 2050, (k) decrease in total pre-frail individuals in 2050, (l) increase in total robust individuals in 2050.

From 2025 to 2050, the Baseline Scenario showed a yearly increase in new frail individuals, rising from 24,558 (95% CrI: 17,809-31,472) to 41,019 (31,127–51,542). All scenarios with interventions reduce new frail individuals, with reductions increasing over time, even in the case of 5% effectiveness. The Both Scenario achieves the largest reduction, reaching 9,013 (6,796–11,895), 4,228 (3,282-5,714), 2,052 (1,749- 2,680), 891 (692-1,210) and 464 (350-603) fewer new frail individuals by 2050 according to 100%, 50%, 25%, 10% and 5% intervention effectiveness, respectively.

All scenarios with interventions increase remaining robust individuals, with yearly growths across all effectiveness. The Both Scenario yields the highest number of remaining robust individuals, exceeding the Baseline Scenario (544,390 (95% CrI: 463,578–643,190) remaining robust individuals) by 78,263 (56,444–98,788), 31,407 (19,834-39,673), 15,793 (10,235-19,562), 6,542 (4,260-8,220), and 3,848 (2,711-4,805) by 2050 across effectiveness scenarios. For individuals reverting to robust, all scenarios show increases over time across all intervention effectiveness. The Both Scenario achieves the largest increase, exceeding the Baseline Scenario (66,936 (64,647-69,899) new robust individuals) by 4,637 (3,065-6,784), 2,933 (1,899-3,931), 1,082 (690-1,565), 618 (397- 849), and 197 (118-292), respectively.

In 2030 and 2050, all scenarios with interventions mitigate frailty by reducing new and total frail individuals while increasing new and total robust individuals across all age groups compared to the Baseline Scenario (Figure 4). The reductions in both new frail individuals and total frail individuals show increasing trends with age in 2050. Among individuals older than 80 years, the Both Scenario achieves the highest reductions by 33.8% (95% CrI: 30.7%-37.1%) and 25.3% (24.9%-26.5%), reaching 2,669 (2,247-2,966) and 26,048 (18,827-34,774) fewer new frail and total frail individuals compared to the Baseline scenario 7,905 (6,063-96,54) new frail individuals and 102,803 (75,669- 131,031) total frail individuals).

### Healthcare Utilization Analysis

The Both Scenario achieves the largest reduction (Figure 5), with 460,914 (95% CrI: 251,312–607,011), 203,836 (143,225-294,561), 115,801 (68,189-159,570), 42,088 (25,211- 66,840), and 21,097 (13,064-30,670) fewer SOCV across the intervention effectiveness scenarios than the Baseline Scenario, where the number of SOCV is projected to be 7,097,602 (5,979,538-8,003,813) by 2050. Similarly, the numbers of EDV show decreases rising annually. By 2050, the Both Scenario reaches the highest reduction with 58,719 (33,156-77,225), 28,110 (19,706-39,115), 14,606 (8,314-21,506), 5,597 (3,384-9,017), and 2,826 (1,662-4,093) fewer EDV, respectively, compared to the Baseline Scenario, which projects 610,145 (499,172-697,157) EDV by 2050. Hospitalizations followed the same trend, with annual decreases growing across all scenarios with interventions. By 2050, the Both Scenario reaches the largest reduction with 64,763 (36,569-85,174), 31,004 (21,735- 43,141), 16,109 (9,170-23,720), 6,174 (3,733-9,945), and 3,117 (1,833-4,514) fewer hospitalizations across the intervention effectiveness scenarios compared to the Baseline Scenario, which projects 472,748 hospitalizations (372,569-549,991).

**Figure 5:**
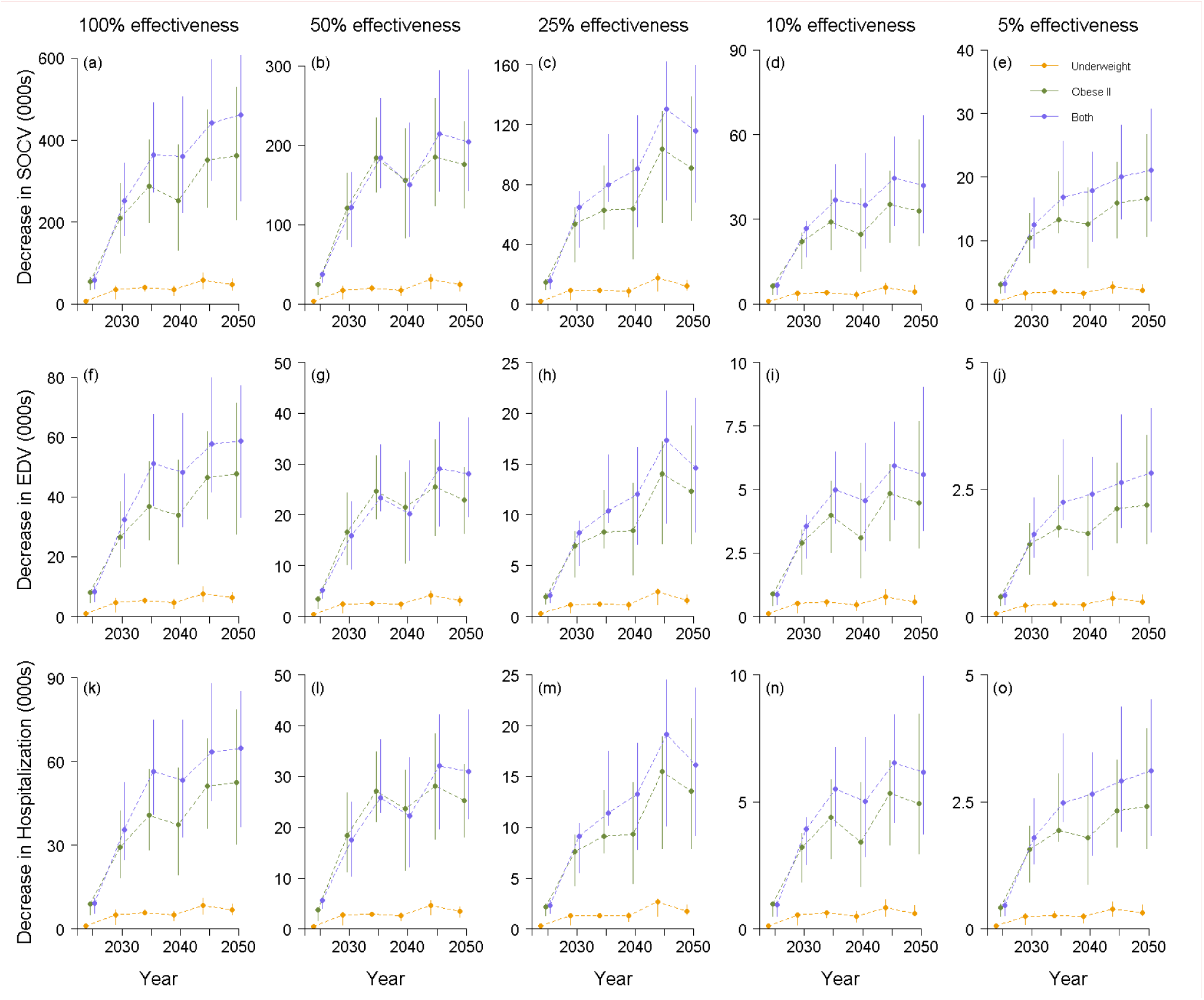
Reduction in healthcare utilization of SOCV (Row 1), EDV (Row 2), and hospitalization (Row 3) in each scenario with weight management interventions (Underweight, Obese II, Both) compared to the Baseline scenario from 2025 to 2050. The five columns represent five levels of weight-control intervention effectiveness, with 100%, 50%, 25%, 10%, and 5% of the obese II individuals shifting to obese I or underweight individuals shifting to the normal weight category. Estimates are presented as **posterior means and 95% credible intervals as error bars**. **SOCV, specialist outpatient clinic visits; EDV, emergency department visits.**

## Discussion

In this study, we examined the impact of widely utilized weight management interventions on pre-frailty and frailty. Our projections indicate significant increases in pre-frailty and frailty by 2030 and 2050, underscoring a three-fold rise in prevalence compared to 2011. We discuss the effectiveness of these strategies in mitigating frailty risks and highlight the implications for public health policies aimed at reducing frailty prevalence and enhancing healthcare outcomes.

Here we created a microsimulation model to forecast pre-frail and frail prevalences in an aging population of 3,353,032 people, exploring the impact of weight management interventions that are widely utilised as primary and secondary measures to avert these outcomes. We estimate that 707,535 (95% CrI: 657,925-743,032) and 125,686 (88,168- 167,322) of all individuals across age, ethnicities and genders will be pre-frail and frail by 2030, increasing up to 836,533 (792,202-860,643) and 201,859 (122,245-263,199) by 2050 respectively, representing a three-fold increase in prevalence compared to 2011. Outside of Southeast Asia, a systematic review of various longitudinal studies from 1987 to 2020 examining frailty trajectories in Netherlands, the USA, Swedish, China, Ireland, UK, Sweden, and other European countries found that frailty prevalence has been increasing^52^. A modelling study in Australia estimated that in 2016, approximately 415,769 people aged 65 years or more were frail and almost 1·7 million people were pre-frail, reaching 609,306 frail and 2,248,977 pre-frail individuals by 2027^53^. In England, simulation projections indicated that frailty will increase by 7·1%, from 41·5% to 48·7% between 2017 and 2027^54^.

Reducing frailty prevalence is a public health priority as frailty results in adverse health outcomes, increased hospitalization rates and an overall decline in the quality of life with higher healthcare utilization. Frailty is potentially reversible therefore identifying and addressing frailty earlier can benefit both individuals and healthcare systems^55^. In a study in Singapore^56^ involving 149 participants, which assessed the effects of six-month nutritional supplementation and cognitive training interventions on frailty reduction, significant improvements were observed at three and six months with benefits persisting at 12 months. Frailty scores and status declined across all intervention groups compared to the control group of usual care, with odds ratios of 2·98 for nutritional supplementation and 2·89 for cognitive training. Frailty prevalence decreased by 16% to 22% in the intervention groups, demonstrating that targeted interventions, particularly the combination approach, effectively mitigate frailty in older adults. Within a 3-month intervention study and four-month follow-up in Japan among 131 frail older women, those assigned to the milk fat globule membrane supplementation and exercise group had a frailty reversal rate of 45·5%^57^.

We found that the prevalence of frailty among underweight and obese II increased from 5·3% and 4·8% in 2011, to 11·2% (95% CrI: 7·9%-14·7%) and 23·1% (17·6%-28·1%) by 2050 respectively. Similar associations between BMI values and frailty prevalence have been observed in other studies. In a cross-sectional study in Japan with 7,191 participants in 2011-2012, the odds ratios for frailty prevalence were 2·04 (1·58-2·63) for the underweight and 1·54 (1·15-2·07) for the obese^43^. Additionally, a study with 4,019 participants from the Netherlands, conducted between 2008-2012, found a statistically significant association between BMI and physical frailty^58^. After adjusting for sex, age, level of education, and smoking status, the U-shaped association between BMI and physical frailty remained, with the prevalence of physical frailty being 8·2% in underweight participants, 2·9% in normal weight participants, 2·6% in overweight participants, and 5·0% in obese participants. Furthermore, a meta-analysis of seven studies involving 23,043 individuals from Chile, Spain, England, America, Finland, and Australia (spanning 1974-2022) explored the relationship between obesity and the risk of frailty. Using a random effects model, the summary odds ratio (OR) showed that adults in the obese BMI category were at a significantly higher risk of frailty compared to those in the normal weight group (OR = 1·74, 95% CI: 1·21-2·51, P<0·01). Similarly, the relationship between underweight and the risk of frailty was examined in four studies with 17,019 individuals from Chile, England, America, and Australia spanning the years 1995-2022. The combined results from the random effects model indicated that older adults in the underweight BMI category had a higher risk of frailty than those in the normal weight group (OR = 1·70, 95% CI: 1·13-2·57, P<0·01)^59^.

In our study, with weight management for both the underweight and obese II groups, we estimated a reduction in prefrailty and frailty by 10,395 (95% CrI: 6,079-13,364; reduction: 6·1% (4.9%-13·2%)) and 53,088 (39,484-63,635; reduction: 19.7% (14·8%- 24.7%)) by 2050. In a randomised controlled trial (RCT) of 27 obese frail volunteers in 2003, after six months of a calorie-restricted diet aimed at creating an energy deficit of about 750 kcal/day, the treatment group lost 8·4% ± 5·6% of body weight and frailty scores where significantly improved when assessed with the Physical Performance Test score (2·6 ± 2·5 vs 0·1 ± 1·0; P<0·05), peak oxygen consumption (1·7 ± 1·6 vs 0·3 ± 1·1 mL/min per kilogram; P<0·05), and Functional Status Questionnaire score (2·9 ± 3·7 vs −0·2 ± 3·9; P<0·05)^60^. In the US between 2012-2014, the MEASUR-UP RCT implemented the intervention of a six-month reduced calorie diet at two protein levels to 67 obese II individuals, improving physical composite and physical functioning scores by 9·2 (P<0·001) and 12·7 (P<0·001) at six months^61^. In France, an RCT across retirement homes found that dietary supplements increased muscle power by 57% at three months (P<0·05) with an average BMI increase of 3·65%^62^. Relatively modest weight changes, such as gaining or losing less than five kg, correspond to the substantial majority of transitions from underweight to normal weight and from obese II to obese I, which are achievable for most individuals through long-term, community and clinical weight management programmes in Singapore^63–65^. Under the purview of Healthier SG, which has enrolled over one million adults aged above 40 into its enhanced screening and GP-patient tracking programme^66^, sustained BMI changes of up to five points are feasible under a long-term national implementation scheme.

Our study supports the need for early identification^67^ and weight management interventions among older individuals to reduce frailty progression rates. A study conducted a CRT from 2015 to 2017 in Taiwan for 319 pre-frail and frail older adults exhibited that at the six-month measurement, the frailty scores significantly improved in both the nutrition group (difference from the baseline: -0.28; 95% CI: -0.41,-0.11) and the combined exercise and nutrition group (-0.34; 95% CI: -0.52,-0.16) compared to the control group^68^. In a study of 738 adults aged 60 and above in Singapore, 90·5% exceeded sugar intake, 68·5% of men and 57·1% of women exceed saturated fat intake, and 49·5% and 55·3% had inadequate dietary calcium intake respectively - 22·3% of older adults were at moderate to high malnutrition risk^69^. Programmes such as the Healthier Dining Programme (HDP)^70^, which increases access to healthier food and drink options by offering grants to food and beverage businesses to promote healthier meal options, supporting many older populations that regularly dine at food stalls, could be further developed to provide more fresh produce, less saturated fat, and more protein within typical meals. Counselling and diet management programmes with general practitioners (GPs) under Healthier SG^71^, an initiative that aims for GPs to follow individuals through their lifetime for long-term physical health monitoring, could also be utilised to encourage weight management among older adults. Among the underweight, additional oral supplementation through subsidised schemes could be utilised as a study of 811 participants aged 65 or over who underwent this intervention had an average of 5% weight gain and increased physical strength^72^. Policy recommendations could additionally focus on personalized nutrition programs with targeted subsidies for nutrient-dense meal plans, particularly benefiting underweight seniors, at food stalls as well as within Meals on Wheels^73^ to target a wider population. Community-based weight management centers could be established to improve access to structured interventions such as dietary education, cooking workshops, and peer-led support programmes. Routine BMI and frailty screenings would also be highly recommended to enable early detection and timely intervention at the robust and at-risk of becoming pre-frail stage. An integrated plan targeting the malnutrition of older adults within the community, workplace, and at home is urgently needed, with necessary healthcare support to maintain robustness and keep individuals at a healthy BMI, which has shown can be an important factor in preventing and averting frailty.

Our research has several limitations. We assumed that the proportions of robust, pre-frail and frail adults and the remission between each status from the Singapore Longitudinal Ageing Study 2 with (SLAS-2) in 2011 and a 5-year follow-up in 2016^10^ were representative of the whole population, where the coverage was five years. We also utilised age, ethnicity, and gender as the main factors in determining frailty risk, as we lacked data on the potential additional risk of other covariates such as smoking, physical activity, dietary habits, and chronic diseases. Our estimates of healthcare utilization do not account for heterogeneity in the impact of frailty across individuals. Although we utilised a data-driven approach, we simplified the model to estimate prevalences of pre-frailty and frailty. Although excess BMI, including extreme obesity, has been associated with increased frailty risk in multiple studies^19^, several additional indirect pathways also exist such as the interaction between diabetes and frailty, which requires further exploration. The bidirectional causality of frailty and comorbidities also merits investigation in additional studies. Finally, our population projections may also not truly reflect future trends as policy changes may alter migration and fertility rates. However, our model’s frailty prevalence estimates align to a large study conducted in 2021 by the Singapore Ministry Health Data Analytics Division from data collected for 425,293 seniors 60 and above (6.9%, 8.2% [5.8%-11.1%])^74^. Data on frailty and pre-frailty estimates is scarce not only in Singapore but in the wider region of the Western Pacific, making modelling studies imperative for evidence-based decision making but such analyses should be repeated with the introduction of new prevalence estimates across time to improve the accuracy of projections.

Despite the limitations, this study developed a microsimulation model using available public data, estimated transition probabilities accounting for key risk factors, and assessed the impact of weight management interventions which are widely implemented globally. National-level projections of elderly pre-frailty and frailty in scenarios with weight management interventions enable the quantification of uncertainties in pre-frail and frail burdens, enhancing the understanding of frailty progressions and the effectiveness of ongoing population-level interventions in reducing pre-frailty, frailty, and associated healthcare utilization in Singapore. Our findings support the recommendation of health policies and services that help older adults shift from underweight or obese BMI categories towards a normal weight, thereby reducing frailty risks in Asian populations. It is also worth highlighting that our model is sufficiently generic to be applied to other aging settings, given the availability of relevant demographic statistics.

## Contributors

Conceptualization, XZ and BD; Investigation, XZ and BD; Formal analysis, XZ and JY; Data visualization, XZ, SJ, and BD; Methodology, XZ, BD, and HS; Project administration, BD; Supervision, BD; Writing – original draft, XZ; Writing – review & editing, All authors (i.e., XZ, SJ, HS, JY, and BD). All authors (i.e., XZ, SJ, HS, JY, and BD) agreed to submit the manuscript.

## Data Sharing Statement

This study does not involve any patient data or participant data. Readers can access the data used in this study from the links to public domain resources provided in the article.

## Declaration of Interests

The authors declare that they have no conflicts of interest.

## Data Availability

All data produced in the present study are available upon reasonable request to the authors.

## Acknowledgements

This research was supported by the Population Health Metrics and Analytics project, the Ministry of Health and National Innovation Challenge (NIC Ageing), Healthy Longevity Catalyst Awards (HLCA) MOH-HLCA22Feb-0007.

## Supplementary Information

### Sample Size

The total sample size of 3,353,032 in our microsimulation model covers individuals aged 55 and above across three main ethnicities—Chinese, Malays and Indians in Singapore from 2011 to 2050. We utilized the Demographic Epidemiological Model of Singapore (DEMOS) to generate individual data, representing each resident in Singapore (including citizens and permanent residents) from 1990 to 2050. Next, we projected the frailty states for each older individual aged 55 and above in DEMOS from 2011 to 2050.

We validated the sample size by comparing the total number of older individuals aged 55 and above in DEMOS with the number of Singapore residents published by the Singapore Department of Statistics^1^ from 2011 to 2024 in Figure S1. The two numbers align, indicating that DEMOS’ modelled estimates reflect the population counts for Singapore.

**Figure S1.**
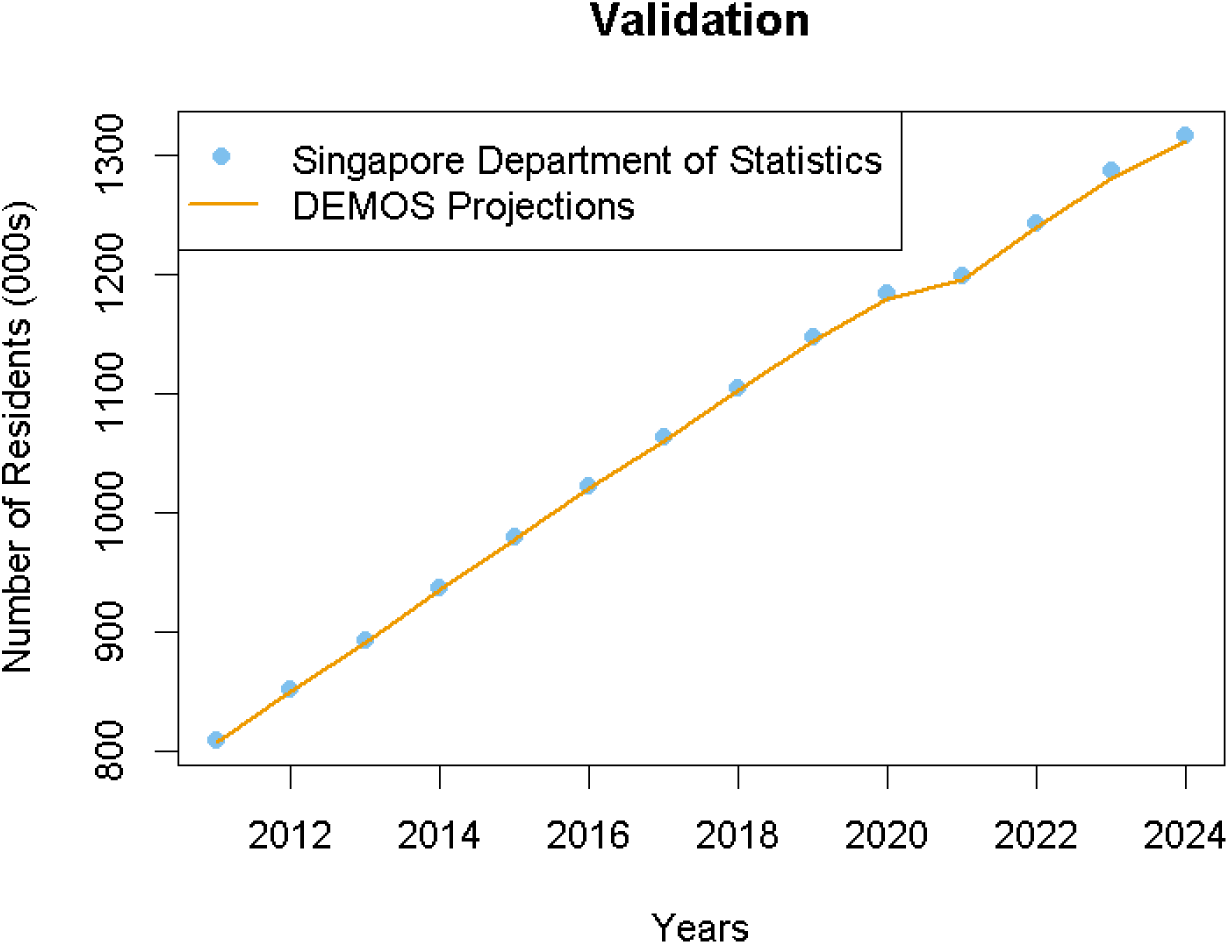
The number of older residents aged 55 and above in Singapore: comparison between Singapore Department of Statistics data and DEMOS data from 2011 to 2024.

### Migration Assumptions

Regarding migration, we assume a migration rate with small variance from 2012 onwards. Using published fertility rates and mortality rates^2,3^, we estimated the yearly migration rates from 1990 to 2020 in Figure S3. For future projections, we calculate the mean and variance of migration rates from the 2010-2020 period, and simulate future migration by applying the mean with variance we calculated previously.

**Figure S2.**
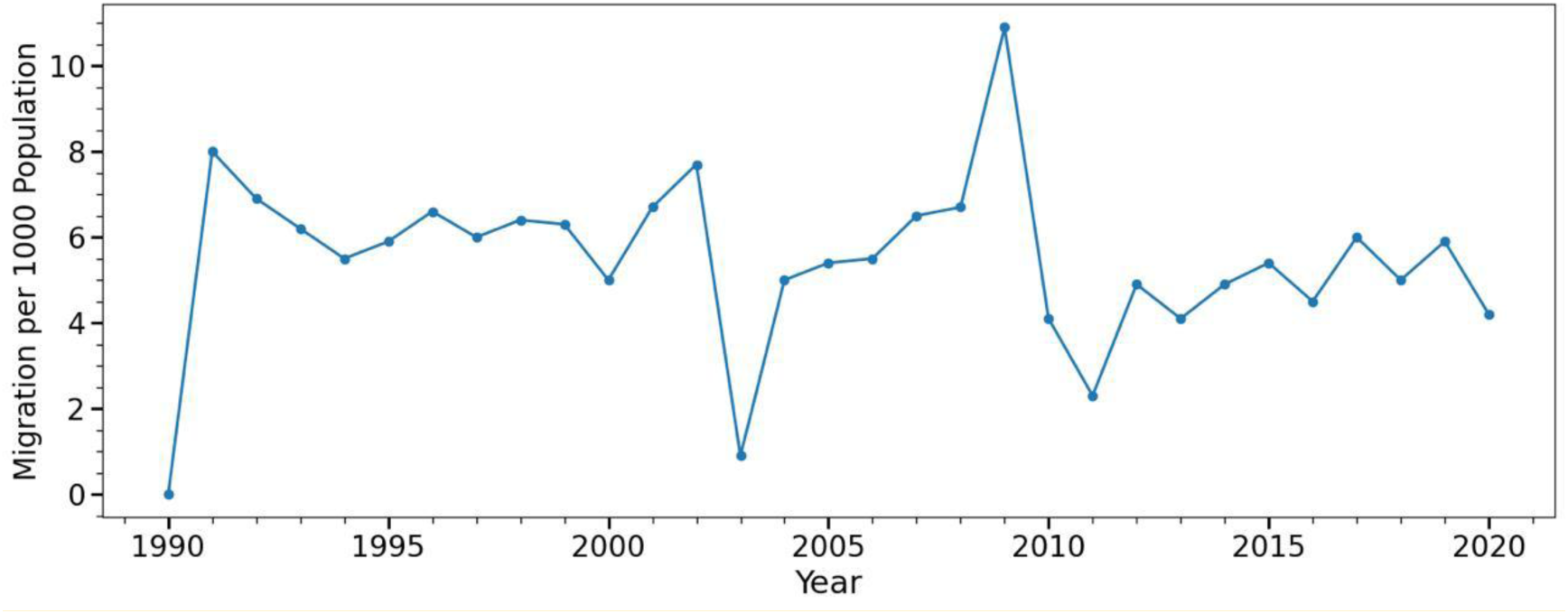
Singapore migration trends (number of migrants per 1000 resident population) for 1990- 2020.

### Fertility Rates Model

We utilised a model framework to forecast total fertility rates which has been described elsewhere^4^. In summary, the three major ethnic groups in Singapore; Chinese (74%), Malays (13%), and Indians (9%), exhibit distinct fertility patterns. For the population not classified within these three ethnic groups, we extrapolated their fertility rates from Malays due to the small sample size of that group and similar observed values.

The data utilized for these models were annual fertility rates for each ethnic group in the ‘low fertility phase’, which began when two consecutive decreases in the fertility rate led to its falling below a threshold of 2.1. The phase spanned from 1975- to 2022 for Chinese and Indians, and from 2003 to 2022 for Malays. Bayesian Structural Time Series (BSTS) models were employed for (long-term) forecasting where a logarithmic transformation was applied to better capture the decelerating rate of decline during this phase.

For each ethnic group, let 𝑌_𝑡_ denote the logarithm of fertility rate at time (year) 𝑡. The BSTS model can be written as

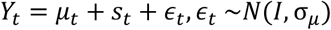

where 𝜇_𝑡_ represents the trend, 𝑠_𝑡_ the seasonality, and 𝜖_𝑡_ the residual. When the ethnic group was Chinese or Indian, a semi-local linear trend model was utilized, represented as

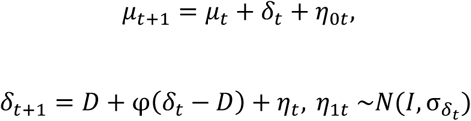

where 𝐷 is the long-term slope of the trend component, towards which 𝛿_𝑡_ will eventually revert, 𝜌 determines the memory in autoregressive (AR) deviations from the long-term trend, and both 𝜂_0𝑡_ and 𝜂_1𝑡_ are random noises. For Malays, however, an AR(1) model was applied due to the relative simple pattern of the time series, i.e.,

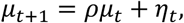

where|𝜌| < 1 and 𝜂_𝑡_ is the random noise. Furthermore, the seasonality parameter, 𝑠_𝑡_ was only non-zero and time-dependent for Chinese, whose fertility rates exhibited a 12-year cyclic pattern.

For each ethnicity, we split the time series into training and validation sets, and simulated 5000 MCMC draws from the model. The means of these draws were utilized as point estimates. The models demonstrated high prediction accuracy across the testing sets (Table S1).

**Table S1:** Model performance, measured by main absolute percentage error (MAPE) for the three BSTS models.

|  | **Training set**<br>(year) | **Validation set**<br>(year) | **MAPE<sub>Train</sub>**<br>(%) | **MAPE<sub>Valid</sub>**<br>(%) |
| --- | --- | --- | --- | --- |
| **Chinese** | 1975–2010 | 2011–2022 | 4.76 | 3.71 |
| **Malays** | 2003–2017 | 2018–2022 | 5.91 | 4.69 |
| **Indians** | 1975–2010 | 2011–2022 | 3.33 | 4.77 |

**Figure S3.**
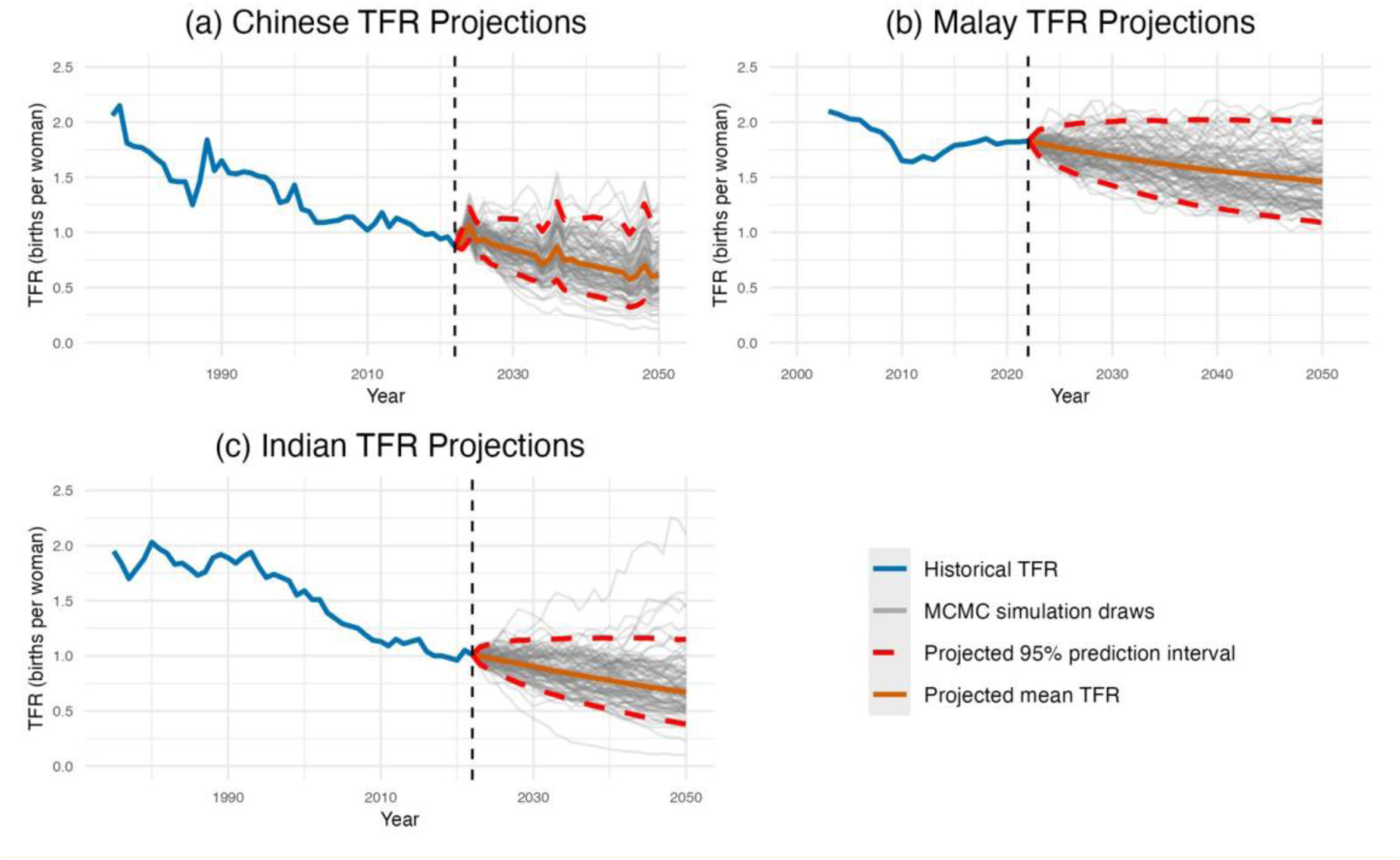
Projected total fertility rates for Chinese (a), Malays (b), and Indians (c) from 2023 to 2050, with grey lines representing 100 of the MCMC simulation draws.

The BSTS model employs Markov Chain Monte Carlo (MCMC) methods for Bayesian inference. Priors are specified as follows: an inverse gamma distribution is placed on the level standard deviation (σ_𝜇_) and the slope standard deviation (σ_𝛿𝑡_), a Gaussian prior governs the long-run slope parameter (D), and the AR(1) coefficient (φ) may adopt a truncated Gaussian prior. To reduce the impact of initial values, the first 10% of MCMC iterations are discarded as burn-in. Convergence of the parameters is assessed by examining trace plots of posterior draws, along with conducting Heidelberger and Welch’s convergence diagnostic tests on the posterior draws in R.

### Bayesian Multistate Model

The original multistate model proposed by Jackson et al. consists of three states, health, disease, and death, and assumes transitions from health to disease, from disease to death, and from disease back to health^5^, with the schematic structure [Health] ↔ [Disease] → [Death]^5^. This model is implemented in the disbayes R package, which uses observed cross-sectional data on disease counts from finite populations to conduct Bayesian inference for age-specific incidence rates, remission rates, case fatality rates, prevalence, and other related parameters. Based on this framework, we developed a Bayesian multistate model to estimate age-specific transition probabilities between robust, pre-frail, and frail states using the disbayes package in R, stratified by gender and ethnicity. Our approach focuses on the progression and remission within adjacent frailty states and excludes mortality or case fatality from the analysis. We adopted a two-stage modelling strategy with the schematic structure [Robust state] ↔ [Pre-frail state] ↔ [Frail state] in Figure S1, using two separate datasets to estimate transition probabilities between robust and pre-frail states, and between pre-frail and frail states.

**Figure S4:**
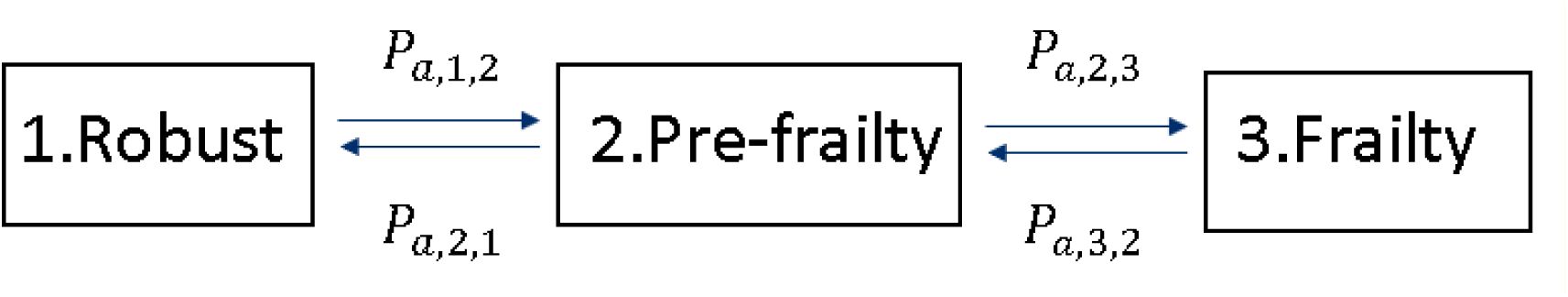
Bayesian multistate model with robust, pre-frail, and frail states and transition probabilities from robust to pre-frail state (*P*_a_,1,2), from pre-frail to robust state (*P*_a_,2,1), from pre-frail to frail state (*P*_a_,2,3), and from frail to pre-frail state (*P*_a_,3,2) for individuals at each age a.

The model assumes each individual may transition only between adjacent states and at most once per year. In the first stage, for each gender and ethnicity stratum, we constructed a dataset including the number of individuals at risk (i.e., robust individuals)(𝑛_a_^(*i*𝑛𝑐,1)^) at each age 𝑎, the number of incident cases of pre-frailty within the next five years (𝑦_a_^(*i*𝑛𝑐,1)^), the number of pre-frail individuals (𝑛_a_^(𝑟𝑒*m*,1)^) at each age 𝑎, and the number of individuals reverting from pre-frail to robust state between 2011 and 2016 (𝑦_a_^(𝑟𝑒*m*,1)^). The dataset was used in disbayes to estimate the five-year transition probabilities from robust to pre-frail state and from pre-frail to robust state. In the second stage, we constructed a dataset focused on pre-frail and frail counts, including the number of pre-frail individuals at each age 𝑎 (𝑛_a_^(*i*𝑛𝑐,2)^), the number of incident cases of frailty (𝑦_a_^(*i*𝑛𝑐,2)^), the number of frail individuals 𝑛_a_^(𝑟𝑒*m*,2)^, and the number of remissions from frail to pre-frail state between 2011 and 2016 (𝑦_a_^(𝑟𝑒*m*,2)^), The second dataset was used to estimate the five-year transition probabilities from pre-frail to frail state, and from frail to pre-frail state.

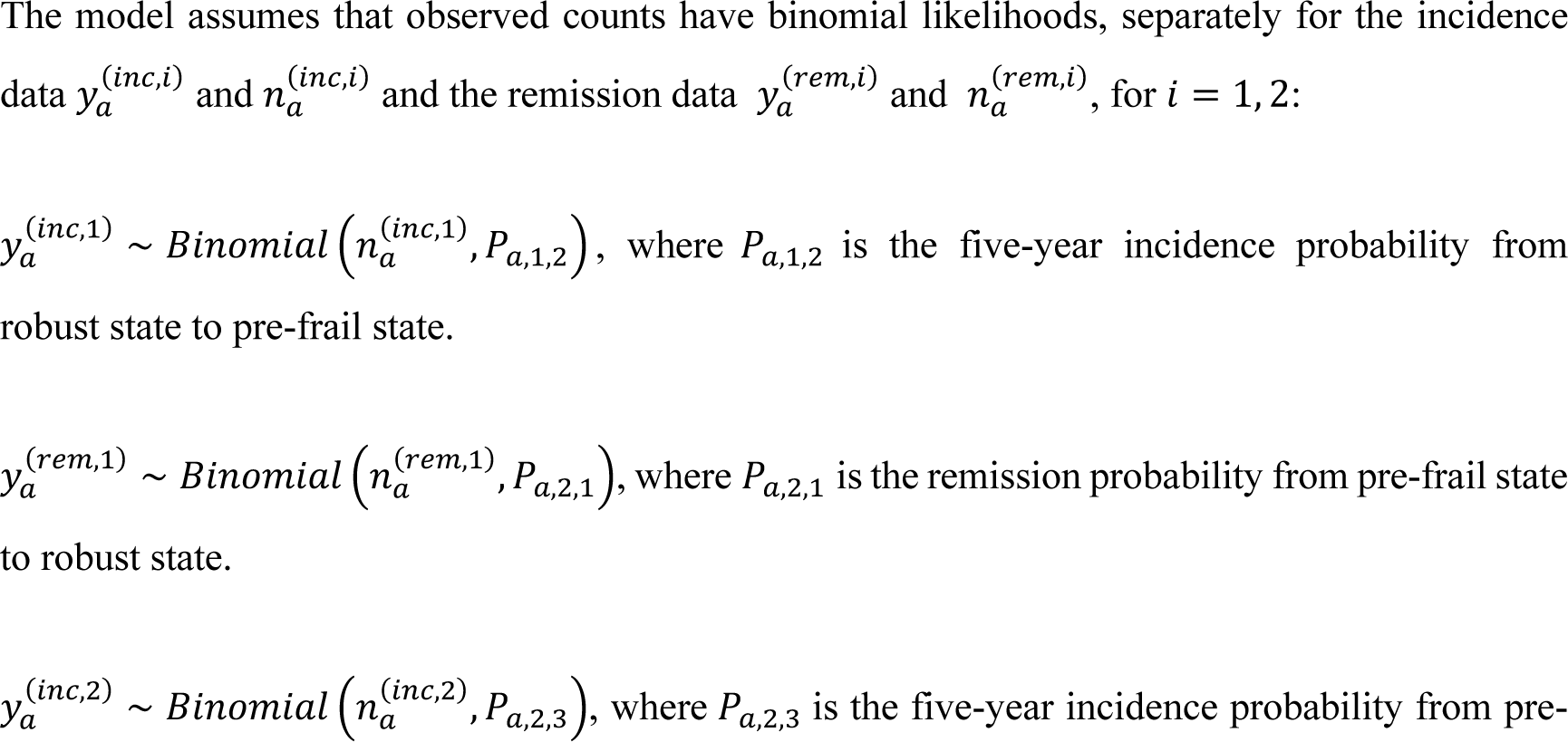
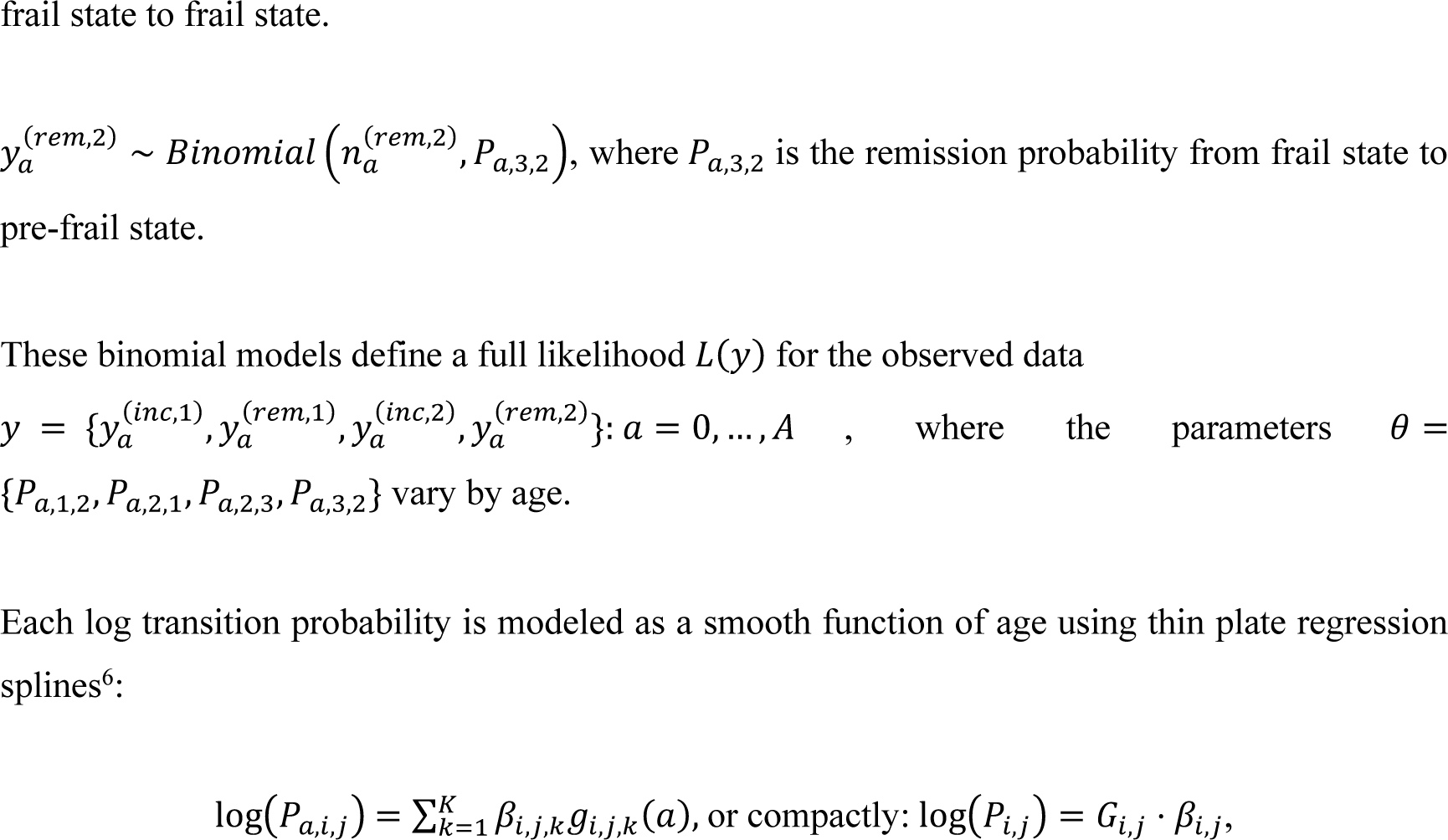

for transitions (*i*, 𝑗) = (1,2), (2,1), (2,3), (3,2) , where 𝑔_*i*,𝑗,𝑘_(𝑎) are spline basis functions, including the intercept term 𝑔_*i*,𝑗,1_(𝑎) = 1, the slope term 𝑔_*i*,𝑗,2_(𝑎) = 𝑎, and the remaining terms 𝑔_*i*,𝑗,𝑘_(𝑎) for 𝑘 ≥ 3 which capture the nonlinear relationship; 𝛽_*i*,𝑗,𝑘_ are coefficients to be estimated, with a common prior distribution 𝛽_*i*,𝑗,𝑘_ ∼ 𝑁(0, 𝜆^2^ ) 𝑓𝑜𝑟 𝑘 ≥ 3. The basic matrix 𝐺_*i*,𝑗_ ∈ 𝑅^𝐴×𝐾^is precomputed, with each row corresponding to an age 𝑎 and each column representing the evaluation of basic function 𝑔_*i*,𝑗,𝑘_(𝑎) for 𝑘 = 1, … , 𝐾 across all ages. The exact form of these basis functions is complex to write explicitly, involving radial basis functions and low-rank approximations derived from thin plate spline theory. They are constructed by applying eigen decomposition to the penalty matrix associated with the spline’s roughness penalty functional, which selects the dominant smooth components of the function space^7^.

Unlike traditional splines, thin plate splines do not use explicit defined knots and manual knots selection is unnecessary. A sufficiently large number of basic spline functions K=10 ensures high flexibility, and the prior distribution 𝑁(0, 𝜆^2^ ) on 𝛽_*i*,𝑗,𝑘_ 𝑓𝑜𝑟 𝑘 ≥ 3 serves as a Bayesian smoothing penalty by shrinking unnecessary terms towards 0, preventing overfitting. The prior standard deviation 𝜆_*i*,𝑗,0_ controls the degree of smoothness, with smaller 𝜆_*i*,𝑗,0_→0 increasing linearity of the curve and large 𝜆_*i*,𝑗,0_ allowing more flexibility. Bayesian updating of 𝜆_*i*,𝑗,0_ enables the model to adapt smoothness to the data. The prior Gamma(2,1) assigned to 𝜆_*i*,𝑗,0_ favors moderate values of 𝜆_*i*,𝑗,0_, resulting in moderate smoothness and preventing extreme shrinkage. For the intercept 𝛽_*i*,𝑗,1_ and slope 𝛽_*i*,𝑗,2_ , weakly informative normal priors 𝑁(0, 100^2^) are assigned to allow the model sufficient flexibility to fit a wide range of plausible baseline levels and trends while avoiding overly extreme values in the absence of data support.

The model is implemented via the disbayes package, which uses the RStan backend to perform Bayesian inference through Hamiltonian Monte Carlo (HMC). Samples of posterior distributions for the spline coefficients 𝛽_*i*,𝑗,𝑘_ from which the age-specific transition probabilities are derived, are generated through the following steps:

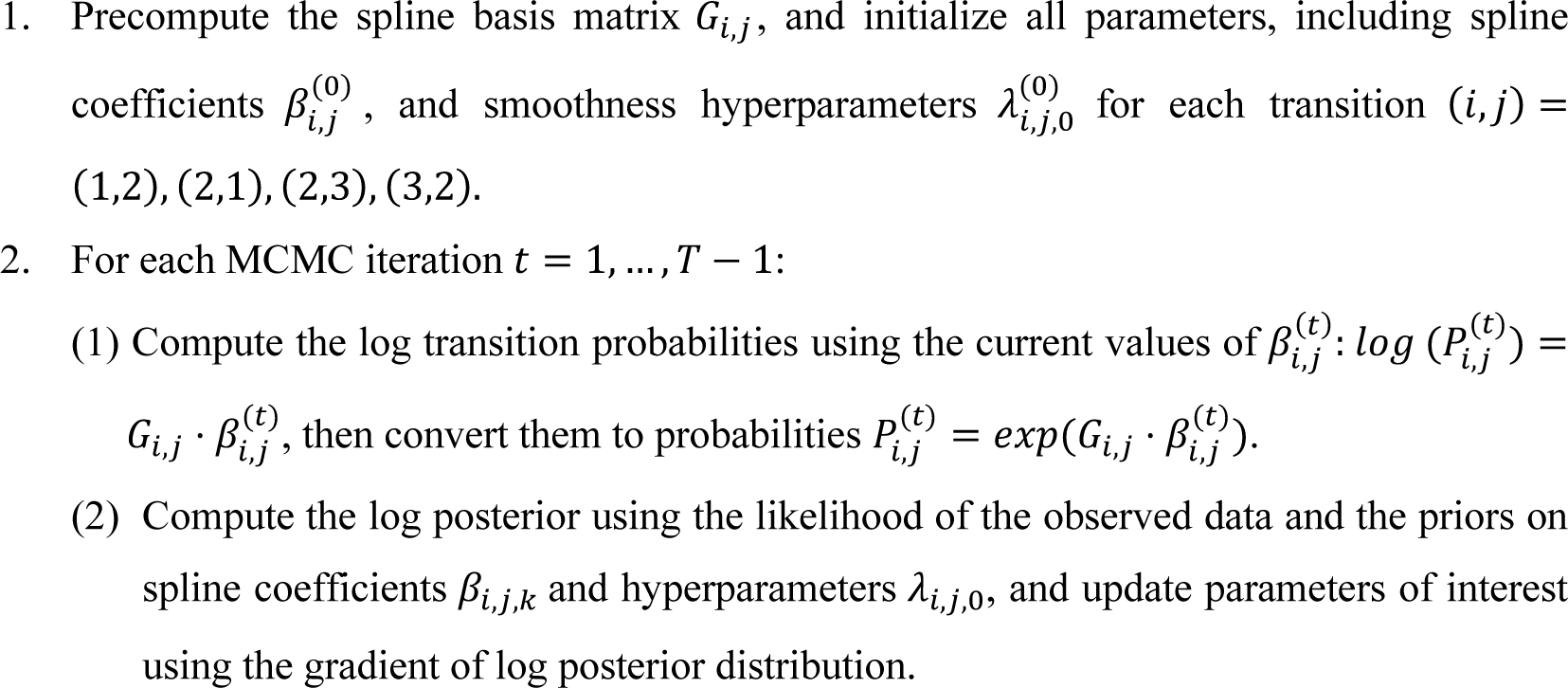

These computations are handled internally by the disbayes package during MCMC sampling, which generates samples from the posterior distribution for parameters including the transition probabilities from the robust state to pre-frailty (*P*_𝑎,1,2_), from pre-frailty to robust state (*P*_𝑎,2,1_). from pre-frailty to frailty (*P*_𝑎,2,3_), and from frailty to pre-frailty (*P*_𝑎,3,2_).

### Risk Factors of Pre-frailty/Frailty Model

Individual-level data containing both frailty states and sociodemographic information (e.g., age, sex, ethnicity, and BMI) were not available to us. Instead, we used a sample of 1,051 community-dwelling older adults in a cohort study, which showed that the prevalence of pre-frailty and frailty significantly increased with age (P<0.001), and differed by gender (P=0.01) and ethnicity (P=0.048)^8^. BMI is also significant (P<0.001), but the distribution of BMI in the sample lacks data with obese II category, which limits the ability to assess the impact of obesity on frailty transitions directly. Therefore, we first estimated transition probabilities considering risk factors of age, gender, and ethnicity, and then incorporated odds ratios of normal weight and other BMI categories on pre-frailty and frail from other studies conducted in Singapore to estimate transition probabilities across BMI categories. This approach allowed us to better reflect the effect of excess BMI, including extreme obesity, which has been shown associated with increased frailty risk in studies^9–13^.

### BMI Intervention Modelling

The means and standard deviations of BMI among robust, pre-frail, and frail older individuals from the sample of 1162 community-dwelling older Singaporeans from SLAS-2 were presented as 24.5 ± 3.5, 23.6 ± 4.3, 22.9 ± 4.5 (mean ± standard deviation), respectively3. Based on the proportions of robust (52.1%), pre-frail (44.6%), and frail (3.29%) individuals in the SLAS-2 sample^14^, we generated three datasets comprising 5,210 robust, 4,460 pre-frail, and 329 frail individuals, respectively. For each dataset, we simulated BMI values using truncated normal distributions, ensuring the means and standard deviations equal to 24.5 and 3.5 for robust individuals, 23.6 and 4.3 for pre-frail individuals, and 22.9 and 4.5 for frail individuals, respectively. We combined the three datasets and analyzed BMI values and frailty states using the logistic regression models to estimate the association of BMI categories including underweight (<18·5 *kg*/*m*^2^), normal weight (18·5–22·9 *kg*/*m*^2^), overweight (23·0–24·9 *kg*/*m*^2^), and obesity I (25·0–29·9 *kg*/*m*^2^) on being pre-frail and frail. Since the proportion of simulated samples with BMI in the Obese II category (≥30.0 kg/m²) was too low to adequately assess its effect on frailty states, we used the observed proportions of robust, pre-frail, and frail individuals within the Obese II category in the sample of 2102 Singapore residents from the Well-being of the Singapore Elderly study (WSES)^15^ to estimate its effect on pre-frailty and frailty compared to normal weight and to calculate the corresponding odds ratios.

To explore the association of BMI categories on pre-frailty and frailty, we fitted two separate logistic regression models using the BMI category as the predictor and frailty status as the binary outcome. The models followed the logistic regression form:

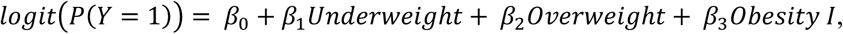

where 𝑌 = 1 represents the outcome of interest, and BMI was treated as a categorical predictor with four levels: underweight (<18·5 *kg*/*m*^2^), normal weight (18·5–22·9 *kg*/*m*^2^; reference category), overweight (23·0–24·9 *kg*/*m*^2^), and obesity I (25·0–29·9 *kg*/*m*^2^).

In the first model, we examined the association between BMI and the likelihood of being pre-frail versus robust. We included only robust and pre-frail individuals in the dataset, coding the outcome variable as 0 for robust and 1 for pre-frail.

In the second model, we assessed the association between BMI and the likelihood of being frail versus pre-frail. This model included only pre-frail and frail individuals, with the outcome coded as 0 for pre-frail and 1 for frail.

### Validation and Optimization

We used grid search, allocating different proportions of robust, pre-frail, and frail individuals for each age, gender, and ethnicity in 2011 and their transitions in 2016 in DEMOS to generate multiple datasets used in our Bayesian multistate model to estimate transition probabilities. Then we used these groups of transition probabilities in DEMOS to project individuals’ frailty states from 2011 to 2016, respectively. We compared the modeled prevalences of pre-frailty and frailty for individuals aged 60 or over with observed prevalences from the SLAS-2 cohort^14^, and chose the group of transition probabilities that minimize the total error. In this way, our estimated prevalence of pre-frailty and frailty (48.9% and 7.5%) matches the observed prevalences

### Estimation of Healthcare Utilization

**Table S2:**
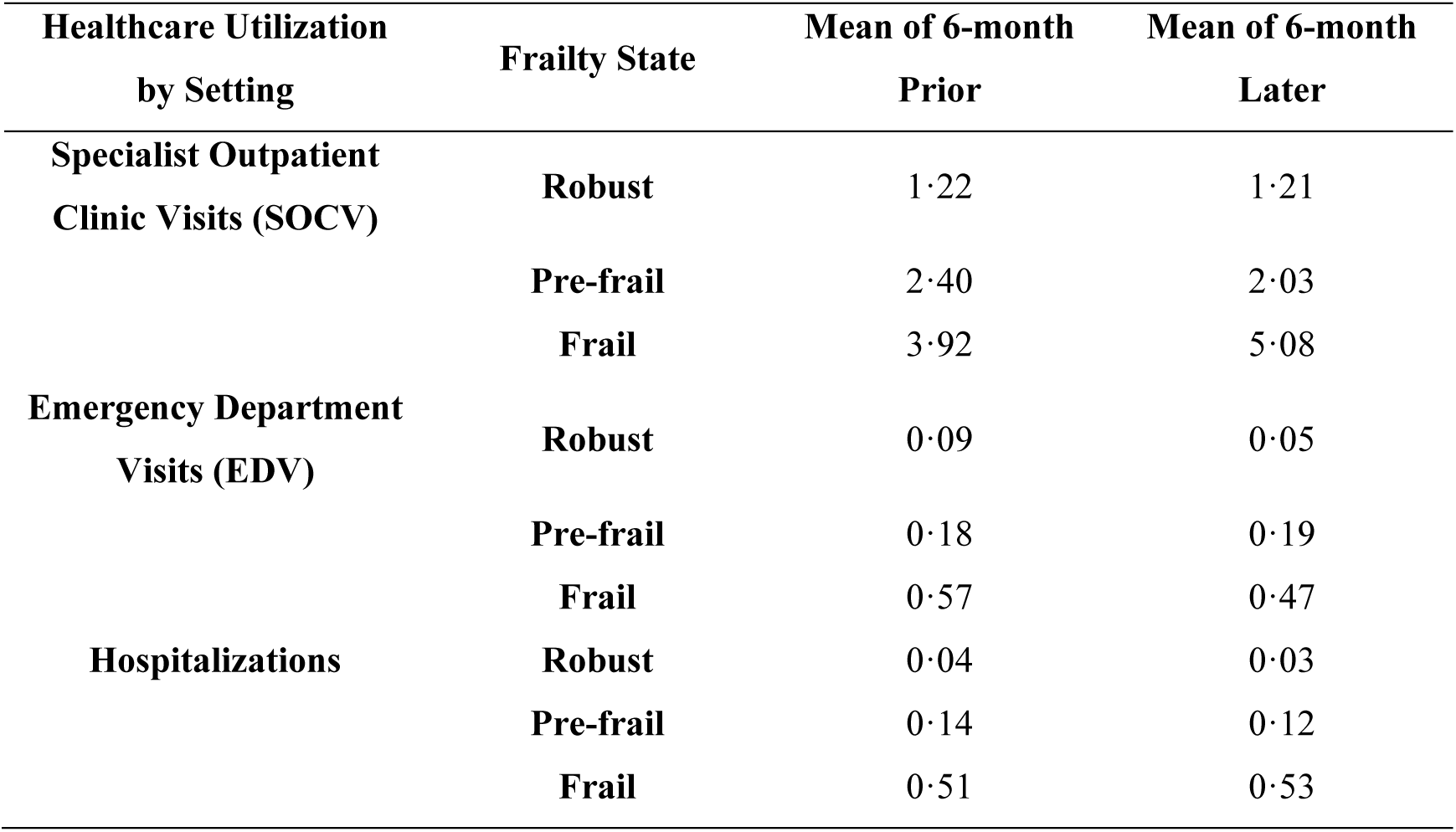
Means of Healthcare Utilization. The mean of 6-month prior and mean of 6-month later are collected from the Population Health Index study covering two periods: six months before and six months after the observation of frailty states4.

| Healthcare Utilization<br>by Setting | Frailty State | Mean of 6-month<br>Prior | Mean of 6-month<br>Later |
| --- | --- | --- | --- |
| Specialist Outpatient<br>Clinic Visits (SOCV) | Robust | 1.22 | 1.21 |
|  | Pre-frail | 2.40 | 2.03 |
|  | Frail | 3.92 | 5.08 |
| Emergency Department<br>Visits (EDV) | Robust | 0.09 | 0.05 |
|  | Pre-frail | 0.18 | 0.19 |
|  | Frail | 0.57 | 0.47 |
| Hospitalizations | Robust | 0.04 | 0.03 |
|  | Pre-frail | 0.14 | 0.12 |
|  | Frail | 0.51 | 0.53 |

### Projection Results across Weight Management Scenarios

**Figure S5:**
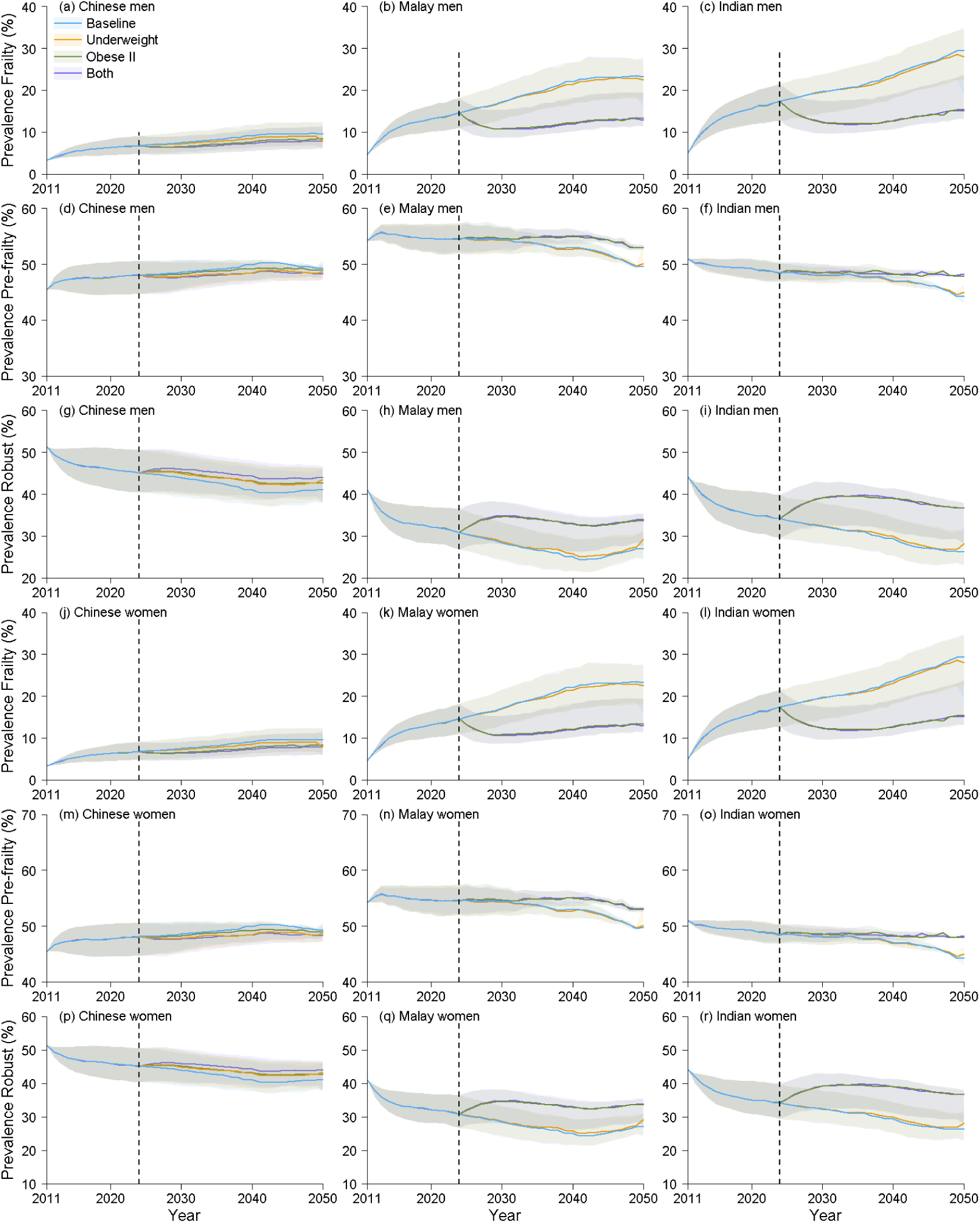
Figure S5. Projected prevalences of the robust, pre-frail, and frail individuals (posterior means and 95% credible intervals as shaded areas) for each ethnicity (Chinese, Indian, and Malay) and gender (women and men) under four scenarios: baseline, underweight to normal weight intervention (“Underweight”) II to obese I intervention (“Obese II”), and the combination of both Underweight and Obese II strategies (“Both”) from 2011 to 2050. (a) frail prevalence for Chinese men, (b) frail prevalence for Malay men, (c) frail prevalence for Indian men, (d) pre-frail prevalence for Chinese men, (e) pre-frail prevalence for Malay men, (f) pre-frail prevalence for Indian men, (g) robust prevalence for Chinese men, (h) robust prevalence for Malay men, (i) robust prevalence for Indian men, (j) frail prevalence for Chinese women, (k) frail prevalence for Malay women, (l) frail prevalence for Indian women, (m) pre-frail prevalence for Chinese women, (n) pre-frail prevalence for Malay women, (o) pre-frail prevalence for Indian women, (p) robust prevalence for Chinese women, (q) robust prevalence for Malay women, (r) robust prevalence for Indian women.

## Notes

### Competing Interest Statement

The authors have declared no competing interest.

### Funding Statement

This study was funded by Singapore Ministry of Health, National Innovation Challenge (NIC Ageing)
Healthy Longevity Catalyst Awards (HLCA).

### Author Declarations

The study used (or will use) ONLY openly available human data that were originally located at: https://pubmed.ncbi.nlm.nih.gov/30498830/ https://pubmed.ncbi.nlm.nih.gov/27576598/ http://www.singstat.gov.sg/find-data/search-by-theme/population/births-and-fertility/latest-data https://pmc.ncbi.nlm.nih.gov/articles/PMC3367999/ http://www.singstat.gov.sg/find-data/search-by-theme/population/death-and-life-expectancy/latest-data https://pubmed.ncbi.nlm.nih.gov/25452860/ https://tablebuilder.singstat.gov.sg/table/TS/M810801 https://hpb.gov.sg/community/national-population-health-survey/survey-findings

### Summary of Updates

Sections on BMI Intervention Modelling and Addition of Frailty-Related Mortality updated to clarify; Figures 2,3,4,5 updated; Section on Results revised; Supplemental files updated.

